# Nutritional, behavioral and anthropometric factors associated with colorectal cancer in Nouakchott, Mauritania: a case–control study

**DOI:** 10.64898/2026.05.23.26353931

**Authors:** Nah Tolba, Adil Najdi, Mohamed El Hfid, Mohameddou Hmeied Maham, Selma Mohmed Brahim, Ahmedou Tolba, Nabila Sellal

**Affiliations:** Health District of Ouad Naga, Ministry of Health, Mauritania; Laboratory of Epidemiology and Public Health, Faculty of Medicine and Pharmacy of Tangier, Abdelmalek Essaâdi University, Tangier, Morocco; Department of Radiotherapy, Mohammed VI University Hospital of Tangier, Faculty of Medicine and Pharmacy of Tangier, Abdelmalek Essaâdi University, Tangier, Morocco; Department of Preventive Medicine, Directorate General of Health Services of the Armed and Security Forces, Ministry of Defence, Veterans Affairs and Martyrs’ Children, Nouakchott, Mauritania; Research Unit, National Oncology Center, Nouakchott, Mauritania; Department of Radiotherapy, Oncology Center, Faculty of Medicine, Pharmacy and Odonto-Stomatology, University of Nouakchott, Nouakchott, Mauritania

**Author notes:** Corresponding author: Dr Nah Tolba, Health District of Ouad Naga Ministry of Health, Mauritania, |. Private Practice in Radiation Oncology, Tangier, Morocco.

**Keywords:** colorectal cancer, risk factors, diet, abdominal obesity, case–control study, Nouakchott, Mauritania

## Abstract

**Background:** Colorectal cancer is a growing public health concern in low- and middle-income countries, particularly in the context of nutritional transition and changing lifestyles. In Mauritania, evidence on factors associated with colorectal cancer remains limited. This study sought to identify nutritional, behavioral and anthropometric factors associated with colorectal cancer among adults living in Nouakchott.

**Methods:** A case–control study was conducted in Nouakchott between January and April 2026. The study included 50 confirmed colorectal cancer cases and 100 controls with no personal history of cancer. Data were collected using a standardized questionnaire covering sociodemographic characteristics, dietary habits, behavioral factors and anthropometric measurements. Crude and adjusted odds ratios with 95% confidence intervals were calculated using binary logistic regression.

**Results:** Low educational level was more frequent among cases than controls, 70.0% versus 27.0%, and remained independently associated with case status after adjustment (aOR = 4.98; 95% CI: 1.81–13.70; p = 0.002). Being married or living with a partner was also associated with case status (aOR = 3.72; 95% CI: 1.19–11.66; p = 0.024). Abdominal obesity was associated with colorectal cancer in bivariate analysis but not after adjustment. High consumption of salty foods remained associated with case status in the multivariate model (aOR = 47.45; 95% CI: 4.83–466.40; p = 0.001). However, this estimate should be interpreted with caution given the wide confidence interval and the limited sample size (n=50 cases). Refined sugars and canned foods were associated with case status only in bivariate analysis. Inverse associations observed for coffee and sugar-sweetened beverages should be interpreted cautiously because of possible reverse causality.

**Conclusion:** Low educational level and high consumption of salty foods were the most defensible factors associated with colorectal cancer in this study. These findings support strengthening nutrition-related prevention and health education interventions in Nouakchott. Larger studies with more detailed dietary assessment are needed to confirm these associations.

## 1 Introduction

Colorectal cancer is ranks among the most common cancers worldwide and represents a growing public health challenge, particularly in low- and middle-income countries. This trend is unfolding against a backdrop of epidemiological and nutritional transition, characterized by urbanization, changes in dietary patterns, reduced physical activity, and increasing rates of overweight and obesity [1,2].

Nutritional, behavioral and anthropometric factors play an important role in the occurrence of colorectal cancer. Epidemiological evidence has highlighted the potential contribution of high consumption of red or processed meat, foods rich in salt, refined sugars or highly processed products, as well as low intake of fruits, vegetables and fiber-rich foods [2,3]. Excess adiposity, especially abdominal obesity, physical inactivity, smoking and some environmental or occupational exposures may also contribute to colorectal carcinogenesis.

In Mauritania, the cancer burden is becoming increasingly concerning. In 2022, estimates reported 3,274 new cancer cases and 2,234 cancer-related deaths in the country [4]. Available data indicate that colorectal cancer is among the cancer sites reported in Mauritania [4]. However, local data specifically addressing nutritional, behavioral and anthropometric factors associated with colorectal cancer remain limited.

Generating context-specific epidemiological data is therefore essential to better understand the profile of colorectal cancer in the Mauritanian population and to guide prevention strategies. This study sought to identify nutritional, behavioral and anthropometric factors associated with colorectal cancer among adults living in Nouakchott, using a case–control design.

This case–control study constitutes the analytical component of a broader doctoral research program on nutritional, behavioral and anthropometric cancer-related risk factors across the three administrative regions of Nouakchott. A complementary cross-sectional study was conducted simultaneously on a representative sample of 1,000 adults from the same population, providing a population-level description of cancer-related risk factor distribution in Nouakchott.

## 2 Methods

### 2.1 Study design, setting and period

This analytical observational case–control study was conducted in Nouakchott, the capital city of Mauritania, from January 25 to April 25, 2026. The study was designed to assess nutritional, behavioral and anthropometric factors associated with colorectal cancer among adults residing in the three administrative regions of Nouakchott: Nouakchott North, Nouakchott West and Nouakchott South.

Cases were recruited from oncology care facilities in Nouakchott, mainly the National Oncology Center, which is the principal national referral facility for cancer diagnosis and treatment in Mauritania. Controls were selected among adults without a personal history of cancer, recruited during the same study period in Nouakchott.

### 2.2 Study population

The study population consisted of adults aged 18 years or older residing in one of the three administrative regions of Nouakchott. Two groups were defined: a case group including patients with confirmed colorectal cancer, and a control group including adults with no personal history of cancer.

Cases were patients with a confirmed diagnosis of colorectal cancer who were receiving care in oncology facilities in Nouakchott. Eligible cases were adults aged 18 years or older, residing in Nouakchott North, Nouakchott West or Nouakchott South, and who agreed to participate in the study after providing informed consent. Patients with another type of cancer, those with incomplete or unusable data, and those who refused to participate were excluded.

Controls were adults with no personal history of cancer, recruited during the same study period in Nouakchott. Eligible controls were aged 18 years or older, resided in one of the three administrative regions of Nouakchott, and provided informed consent. Efforts were made to select controls comparable to cases in terms of age group, sex and region of residence. Individuals with a known history of cancer, those with severe chronic conditions likely to substantially modify dietary habits or anthropometric status, those unable to complete the questionnaire properly, and those who refused to participate were excluded.

### 2.3 Sample size and sampling

The sample size was estimated using Epi Info™, through the module dedicated to case–control studies. The initial calculation was based on a case-to-control ratio of 1:2, a 95% confidence level, an alpha error of 5%, and an expected odds ratio of 2.

Given the limited availability of colorectal cancer cases during the study period and the operational constraints related to recruitment, a feasible sample size was retained. The study therefore included 50 confirmed colorectal cancer cases and 100 controls, for a total of 150 participants. The use of two controls per case was intended to improve the precision of estimates within the available resources.

Cases were recruited consecutively from oncology care facilities in Nouakchott, mainly the National Oncology Center, after verification of their diagnosis from medical records. Controls were selected during the same study period among adults without a personal history of cancer. They were recruited in Nouakchott through non-oncology health service contacts, general consultations, screening activities and community-based contacts. Efforts were made to ensure comparability between cases and controls according to age group, sex and region of residence, in order to reduce selection bias.

The limited sample size (50 cases) may have affected the stability of some estimates in multivariate analysis, particularly for variables with low cell counts. The rule of 10 events per variable (EPV) was not fully met for all variables included in the logistic regression model, which may have led to overfitting and imprecise estimates. This limitation should be considered when interpreting the adjusted odds ratios.

### 2.4 Data collection

Data were collected using a standardized questionnaire initially developed for the cross-sectional phase of the study and subsequently adapted for the case–control component. The questionnaire covered sociodemographic characteristics, dietary habits, behavioral factors, environmental and occupational exposures, physical activity, and anthropometric measurements.

Data collection was performed using the KoboCollect application, which allowed direct, standardized and electronic data entry. The database was then exported to Microsoft Excel for data checking, cleaning and preparation before statistical analysis. Anthropometric measurements, including weight, height, waist circumference and hip circumference, were taken by trained interviewers according to standardized procedures.

### 2.5 Study variables

The dependent variable was case–control status, coded as colorectal cancer case or control. Independent variables included sociodemographic, behavioral, dietary and anthropometric factors. Sociodemographic variables included age, sex, educational level, marital status, occupational level and region of residence.

Behavioral variables included physical activity, smoking status, alcohol consumption, occupational exposure to chemicals, radiation or other potentially harmful agents, sun exposure, frequency of eating outside the home, and frequency of consuming home-prepared meals.

Dietary variables included the consumption of red meat, processed meat, fish, fresh fruits, fresh vegetables, dairy products, refined cereals, refined sugars, salty foods, fried foods, sugar-sweetened beverages, canned foods, tea and coffee. These variables were recoded into analytical categories according to frequency of consumption, mainly as low or high consumption.

Anthropometric variables included weight, height, body mass index, waist circumference and waist-to-hip ratio. Body mass index was classified into underweight (< 18.5 kg/m²), normal weight (18.5–24.9 kg/m²), overweight (25.0–29.9 kg/m²) and obesity (≥ 30.0 kg/m²) according to WHO standard categories. Abdominal obesity was defined using sex-specific waist circumference cut-off points recommended by the World Health Organization: ≥ 88 cm in women and ≥ 102 cm in men.

### 2.6 Data management and statistical analysis

Data were exported to Microsoft Excel for checking and cleaning. This process included verification of internal consistency, identification of outliers, management of missing data and removal of duplicates. Variables were then coded and recoded according to the analytical plan, particularly dietary, anthropometric and behavioral variables. The final dataset was imported into IBM SPSS Statistics, version 22 (IBM Corp., Armonk, NY, USA) for analysis.

Descriptive analysis was performed to summarize the characteristics of cases and controls. Categorical variables were presented as frequencies and percentages.

Bivariate analysis was conducted to assess associations between explanatory variables and case–control status. Proportions were compared using Pearson’s Chi-square test or Fisher’s exact test when Chi-square assumptions were not met. Crude odds ratios and their 95% confidence intervals were calculated for each variable.

Variables associated with colorectal cancer at a p < 0.20 in bivariate analysis, as well as variables considered biologically or epidemiologically relevant, were considered for multivariate analysis. Binary logistic regression was used to identify factors independently associated with colorectal cancer. Age and sex were included in the model as adjustment variables. Results were presented as adjusted odds ratios with 95% confidence intervals and p-values. Statistical significance was set at p < 0.05.

No substantial missing data were observed for the variables included in the final analysis.

### 2.7 Ethical considerations

This study was conducted in accordance with the ethical principles governing research involving human participants and in line with the Declaration of Helsinki. The study protocol received ethical approval from the Health Research Ethics Committee of the Ministry of Health of Mauritania, under reference number 047-2026/MS/CNERS, dated January 22, 2026. The required administrative authorizations were also obtained from the relevant institutions, particularly the National Oncology Center.

Participation was voluntary. Before inclusion, all participants were informed about the study objectives, procedures and intended use of the collected data. Oral informed consent was obtained from each participant, as the study involved only questionnaire administration and anthropometric measurements with no invasive procedures; the oral consent procedure was approved by the ethics committee. Participants were free to refuse participation or withdraw from the study at any time, without any consequences for their care.

Confidentiality and anonymity were ensured throughout the study. Data were coded without direct personal identifiers and used exclusively for scientific purposes.

## 3 Results

### 3.1 General characteristics of participants

A total of 150 participants were included, comprising 50 colorectal cancer cases and 100 controls. The study population included 63 men (42.0%) and 87 women (58.0%), with equal sex distribution across groups.

Most participants were aged 45 years or older, representing 88.0% of cases and 92.0% of controls. The geographic distribution was broadly comparable between groups, with a predominance of residents from Nouakchott North among both cases and controls.

Educational level differed substantially between groups: 54.0% of cases had no formal education, compared with 16.0% of controls, whereas higher education was more frequent among controls than among cases, 56.0% and 22.0%, respectively. The proportion of participants who were married or living with a partner was also higher among cases than among controls, 80.0% versus 53.0%. Occupational level was relatively similar between the two groups.

**Table 1.** General characteristics of participants according to case–control status in the colorectal cancer study.

| Variable | Category | Cases n (%) | Controls n (%) | Total n (%) |
| --- | --- | --- | --- | --- |
| **Sex** | Male | 21 (42.0) | 42 (42.0) | 63 (42.0) |
|  | Female | 29 (58.0) | 58 (58.0) | 87 (58.0) |
| **Age** | < 45 years | 6 (12.0) | 8 (8.0) | 14 (9.3) |
|  | 45–54 years | 16 (32.0) | 27 (27.0) | 43 (28.7) |
|  | 55–64 years | 12 (24.0) | 45 (45.0) | 57 (38.0) |
|  | ≥ 65 years | 16 (32.0) | 20 (20.0) | 36 (24.0) |
| **Region of residence** | Nouakchott North | 30 (60.0) | 56 (56.0) | 86 (57.3) |
|  | Nouakchott West | 9 (18.0) | 20 (20.0) | 29 (19.3) |
|  | Nouakchott South | 11 (22.0) | 24 (24.0) | 35 (23.3) |
| **Educational level** | No formal education | 27 (54.0) | 16 (16.0) | 43 (28.7) |
|  | Primary | 8 (16.0) | 11 (11.0) | 19 (12.7) |
|  | Secondary | 4 (8.0) | 17 (17.0) | 21 (14.0) |
|  | Higher education | 11 (22.0) | 56 (56.0) | 67 (44.7) |
| **Marital status** | Not married | 10 (20.0) | 47 (47.0) | 57 (38.0) |
|  | Married | 40 (80.0) | 53 (53.0) | 93 (62.0) |
| **Occupation** | Unemployed | 22 (44.0) | 47 (47.0) | 69 (46.0) |
|  | Unskilled employment | 11 (22.0) | 28 (28.0) | 39 (26.0) |
|  | Skilled employment | 17 (34.0) | 25 (25.0) | 42 (28.0) |

### 3.2 Bivariate analysis of factors associated with colorectal cancer

#### 3.2.1 Sociodemographic, anthropometric and behavioral factors

In bivariate analysis, low educational level was strongly associated with colorectal cancer. Participants with a low educational level accounted for 70.0% of cases compared with 27.0% of controls and had higher odds of being a case than participants with a higher educational level (crude OR = 6.31; 95% CI: 2.98–13.34; p < 0.001). Marital status was also associated with case status, with married participants or those living with a partner representing 80.0% of cases compared with 53.0% of controls (crude OR = 3.55; 95% CI: 1.61–7.82; p = 0.001).

Among anthropometric factors, two significant associations were observed. Overweight or obesity according to body mass index was less frequent among cases than among controls (34.0% versus 63.0%; crude OR = 0.30; 95% CI: 0.14–0.63; p = 0.009). Conversely, abdominal obesity was more frequent among cases than controls (44.0% versus 27.0%) and was associated with higher odds of being a case (crude OR = 2.12; 95% CI: 1.04–4.33; p = 0.036).

No statistically significant association was observed with age, sex, smoking, physical activity, occupational exposure, eating outside the home, consumption of home-prepared meals, or sun exposure. Alcohol consumption could not be analyzed, as no participant reported alcohol use.

**Table 2.** Bivariate analysis of sociodemographic, anthropometric and behavioral factors associated with colorectal cancer.

| **Variable** | **Category** | **Cases n (%)** | **Controls n (%)** | **Crude OR (95% CI)** | **p-value** |
| --- | --- | --- | --- | --- | --- |
| **Age** | < 45 years | 6 (12.0) | 8 (8.0) | 1.00 ref |  |
|  | ≥ 45 years | 44 (88.0) | 92 (92.0) | 0.64 (0.20–2.04) | 0.450 |
| **Sex** | Male | 21 (42.0) | 42 (42.0) | 1.00 ref |  |
|  | Female | 29 (58.0) | 58 (58.0) | 1.00 (0.51–1.96) | 1.000 |
| **Educational level** | Higher | 15 (30.0) | 73 (73.0) | 1.00 ref |  |
|  | Low | 35 (70.0) | 27 (27.0) | 6.31 (2.98–13.34) | <0.001 |
| **Marital status** | Not married/not living with a partner | 10 (20.0) | 47 (47.0) | 1.00 ref |  |
|  | Married/living with a partner | 40 (80.0) | 53 (53.0) | 3.55 (1.61–7.82) | 0.001 |
| **BMI** | Normal/underweight | 33 (66.0) | 37 (37.0) | 1.00 ref |  |
|  | Overweight/obesity | 17 (34.0) | 63 (63.0) | 0.30 (0.14–0.63) | 0.009 |
| **Abdominal obesity** | No | 28 (56.0) | 73 (73.0) | 1.00 ref |  |
|  | Yes | 22 (44.0) | 27 (27.0) | 2.12 (1.04–4.33) | 0.036 |
| **Smoking** | No | 45 (90.0) | 89 (89.0) | 1.00 ref |  |
|  | Yes | 5 (10.0) | 11 (11.0) | 0.90 (0.29–2.80) | 0.860 |
| **Physical activity** | ≥ 1 time/week | 18 (36.0) | 43 (43.0) | 1.00 ref |  |
|  | None | 32 (64.0) | 57 (57.0) | 1.34 (0.67–2.68) | 0.390 |
| **Occupational exposure** | No | 45 (90.0) | 91 (91.0) | 1.00 ref |  |
|  | Yes | 5 (10.0) | 9 (9.0) | 1.12 (0.34–3.65) | 0.850 |
| **Alcohol consumption** | No | 50<br>(100.0) | 100<br>(100.0) | — | — |
|  | Yes | 0 (0.0) | 0 (0.0) | — | — |
| **Eating outside the home** | Never | 20 (40.0) | 39 (39.0) | 1.00 ref |  |
|  | ≥ 1 time/week | 30 (60.0) | 61 (61.0) | 0.96 (0.48–1.91) | 0.910 |
| **Home-prepared meals** | Daily | 45 (90.0) | 92 (92.0) | 1.00 ref |  |
|  | Not daily | 5 (10.0) | 8 (8.0) | 1.28 (0.39–4.18) | 0.680 |
| **Sun exposure** | None | 35 (70.0) | 65 (65.0) | 1.00 ref |  |
|  | ≥ 1 time/week | 15 (30.0) | 35 (35.0) | 0.80 (0.38–1.66) | 0.550 |

#### 3.2.2 Nutritional factors

In bivariate analysis, several nutritional factors were associated with colorectal cancer. High consumption of refined sugars was more frequent among cases than controls (90.0% versus 63.0%) and was significantly associated with case status (crude OR = 5.29; 95% CI: 1.93–14.49; p = 0.001). High consumption of salty foods was also more frequent among cases (80.0%) than controls (55.0%) and showed a significant association (crude OR = 3.27; 95% CI: 1.44–7.41; p = 0.004). Similarly, high consumption of canned foods was reported by 60.0% of cases compared with 35.0% of controls (crude OR = 2.79; 95% CI: 1.37–5.66; p = 0.004).

Some significant inverse associations were also observed. High consumption of refined cereals was less frequent among cases than controls (30.0% versus 85.0%; crude OR = 0.08; 95% CI: 0.03–0.17; p < 0.001). Likewise, high consumption of sugar-sweetened beverages was reported by 4.0% of cases compared with 32.0% of controls (crude OR = 0.09; 95% CI: 0.02–0.38; p < 0.001). Coffee consumption was also less frequent among cases than controls (4.0% versus 45.0%) and was inversely associated with case status (crude OR = 0.05; 95% CI: 0.01–0.22; p < 0.001).

Non-significant trends were observed for high consumption of processed meat, low consumption of fresh fruits and low consumption of dairy products. No statistically significant association was observed for red meat, fish, fresh vegetables, tea or fried foods.

**Table 3.** Bivariate analysis of nutritional factors associated with colorectal cancer.

| Variable | Category | Cases n (%) | Controls n (%) | Crude OR (95% CI) | p-value |
| --- | --- | --- | --- | --- | --- |
| **Red meat** | < 3 times/week | 12 (24.0) | 29 (29.0) | 1.00 ref |  |
|  | ≥ 3 times/week | 38 (76.0) | 71 (71.0) | 1.29 (0.58–2.86) | 0.530 |
| **Processed meat** | < 3 times/week | 40 (80.0) | 91 (91.0) | 1.00 ref |  |
|  | ≥ 3 times/week | 10 (20.0) | 9 (9.0) | 2.53 (0.96–6.66) | 0.060 |
| **Fish** | < 3 times/week | 42 (84.0) | 76 (76.0) | 1.00 ref |  |
|  | ≥ 3 times/week | 8 (16.0) | 24 (24.0) | 0.60 (0.25–1.44) | 0.250 |
| **Fresh fruits** | ≥ 3 times/week | 15 (30.0) | 45 (45.0) | 1.00 ref |  |
|  | < 3 times/week | 35 (70.0) | 55 (55.0) | 1.91 (0.94–3.86) | 0.070 |
| **Fresh vegetables** | ≥ 3 times/week | 30 (60.0) | 65 (65.0) | 1.00 ref |  |
|  | < 3 times/week | 20 (40.0) | 35 (35.0) | 1.24 (0.62–2.47) | 0.540 |
| **Dairy products** | ≥ 3 times/week | 28 (56.0) | 70 (70.0) | 1.00 ref |  |
|  | < 3 times/week | 22 (44.0) | 30 (30.0) | 1.83 (0.89–3.75) | 0.090 |
| **Refined cereals** | Low | 35 (70.0) | 15 (15.0) | 1.00 ref |  |
|  | High | 15 (30.0) | 85 (85.0) | 0.08 (0.03–0.17) | <0.001 |
| **Refined sugars** | Low | 5 (10.0) | 37 (37.0) | 1.00 ref |  |
|  | High | 45 (90.0) | 63 (63.0) | 5.29 (1.93–14.49) | 0.001 |
| **Salty foods** | < 3 times/week | 10 (20.0) | 45 (45.0) | 1.00 ref |  |
|  | ≥ 3 times/week | 40 (80.0) | 55 (55.0) | 3.27 (1.44–7.41) | 0.004 |
| **Canned foods** | < 3 times/week | 20 (40.0) | 65 (65.0) | 1.00 ref |  |
|  | ≥ 3 times/week | 30 (60.0) | 35 (35.0) | 2.79 (1.37–5.66) | 0.004 |
| **Sugar-sweetened beverages** | Low | 48 (96.0) | 68 (68.0) | 1.00 ref |  |
|  | High | 2 (4.0) | 32 (32.0) | 0.09 (0.02–0.38) | <0.001 |
| **Fried foods** | Low | 45 (90.0) | 80 (80.0) | 1.00 ref |  |
|  | High | 5 (10.0) | 20 (20.0) | 0.44 (0.15–1.28) | 0.121 |
| **Tea** | Low | 12 (24.0) | 26 (26.0) | 1.00 ref |  |
|  | High | 38 (76.0) | 74 (74.0) | 1.11 (0.50–2.49) | 0.791 |
| **Coffee** | Never | 48 (96.0) | 55 (55.0) | 1.00 ref |  |
|  | Consumer | 2 (4.0) | 45 (45.0) | 0.05 (0.01–0.22) | <0.001 |

### 3.3 Multivariate analysis of factors associated with colorectal cancer

Binary logistic regression was performed to identify factors independently associated with colorectal cancer. The model included age, sex, marital status, educational level, abdominal obesity, high consumption of salty foods, coffee consumption, and high consumption of sugar-sweetened beverages.

After adjustment, low educational level remained significantly associated with case status (aOR = 4.98; 95% CI: 1.81–13.70; p = 0.002). Being married or living with a partner was also associated with colorectal cancer (aOR = 3.72; 95% CI: 1.19–11.66; p = 0.024).

High consumption of salty foods remained strongly associated with case status after adjustment (aOR = 47.45; 95% CI: 4.83-466.40; p = 0.001). However, this estimate should be interpreted with great caution. The very wide confidence interval reflects statistical instability related to the limited sample size and the small number of participants in some cells. The rule of 10 events per variable (EPV) was not fully satisfied in our model, which may have led to overfitting. This result should therefore be considered an exploratory signal suggesting an unfavorable salty and processed dietary pattern, rather than evidence of a direct causal effect of salt intake on colorectal cancer risk.

Conversely, coffee consumption (aOR = 0.09; 95% CI: 0.02–0.47; p = 0.004) and high consumption of sugar-sweetened beverages (aOR = 0.11; 95% CI: 0.02–0.59; p = 0.010) were inversely associated with case status. These inverse associations should be considered exploratory, given the potential for reverse causality and changes in dietary habits after diagnosis.

Age, sex and abdominal obesity were not significantly associated with colorectal cancer in the final model.

**Table 4.**
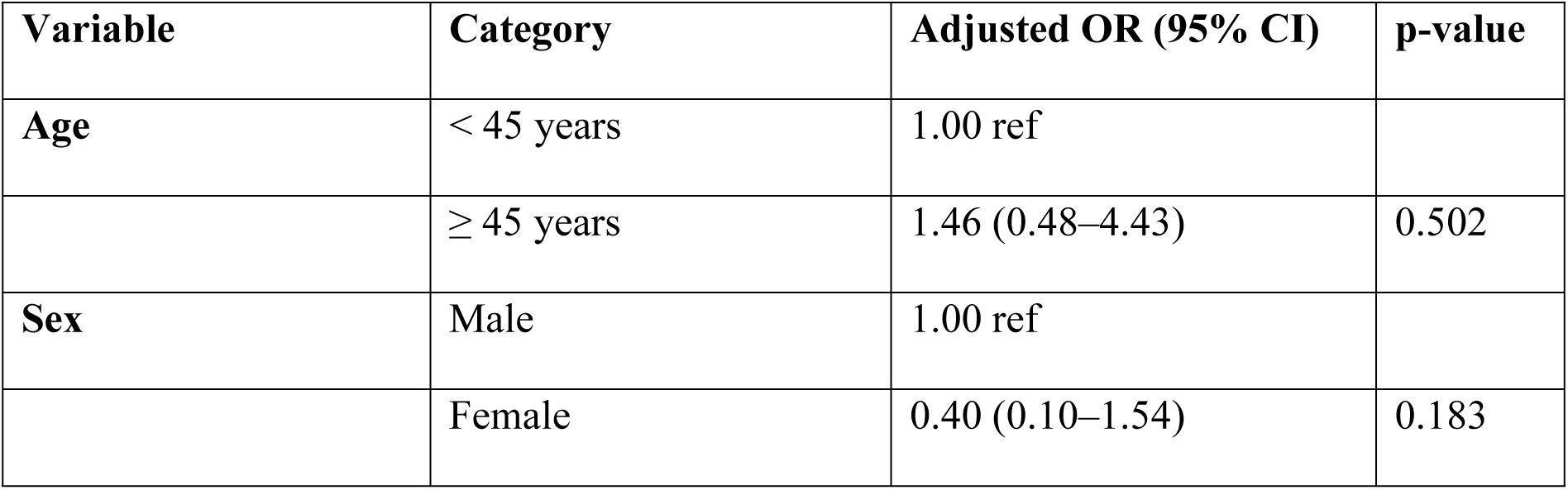
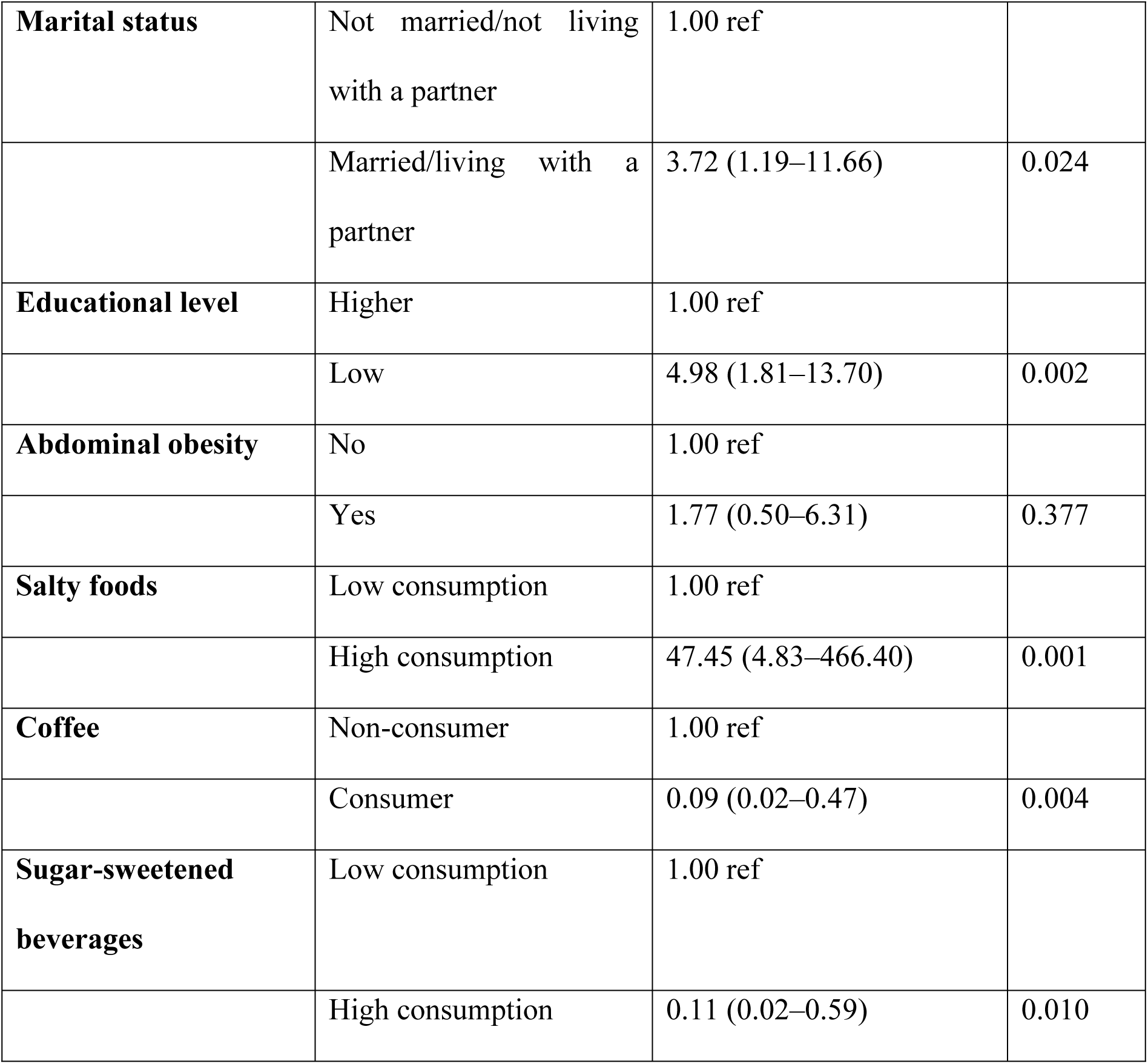
Multivariate model of factors associated with colorectal cancer.

## 4 Discussion

In the present study, cases and controls were broadly comparable in terms of age and sex. Participants aged 45 years or older represented 88.0% of cases and 92.0% of controls, with no significant association with colorectal cancer in either bivariate analysis (crude OR = 0.64; 95% CI: 0.20–2.04; p = 0.450) or multivariate analysis (aOR = 1.46; 95% CI: 0.48–4.43; p = 0.502). Similarly, sex distribution was identical in both groups, with 42.0% men and 58.0% women, and no significant association after adjustment.

These findings differ from those generally reported in the literature. In Uganda, Wismayer et al. conducted a case–control study in 2024 including 128 cases and 256 age- and sex-matched controls; the mean age of participants was 53.5 ± 16.2 years, with a male-to-female ratio of 1:1 [5]. In Algeria, Negrichi and Taleb conducted a case–control study in eastern Algeria and reported the involvement of hereditary, environmental and dietary factors in colorectal cancer risk [6]. In Saudi Arabia, Azzeh et al. also conducted a case–control study in the Makkah region, showing that colorectal cancer mainly affected middle-aged and older adults [7]. At the international level, Keum and Giovannucci emphasized that colorectal cancer incidence increases markedly with age, particularly after 50 years [2].

The absence of an association with age and sex in the present study is likely explained by the relative comparability of cases and controls, the small number of younger participants, and the limited sample size. Therefore, this finding does not challenge the established role of age and sex in the epidemiology of colorectal cancer.

The most important sociodemographic finding concerned educational level. In the present study, low educational level was observed in 70.0% of cases compared with 27.0% of controls. It was strongly associated with colorectal cancer in bivariate analysis (crude OR = 6.31; 95% CI: 2.98–13.34; p < 0.001), and this association persisted after adjustment (aOR = 4.98; 95% CI: 1.81–13.70; p = 0.002). This finding is consistent with the study by Negrichi and Taleb in Algeria, which reported that higher educational level was associated with a lower risk of colorectal cancer [6]. It is also in line with international data from Doubeni et al. in the United States in 2012, who showed that socioeconomic differences in colorectal cancer incidence were partly explained by health behaviors and obesity, with behavioral factors and BMI accounting for an estimated 43.9% of the association related to educational level [8].

In the context of Nouakchott, low educational level should be interpreted as a marker of social and health vulnerability. It may influence dietary choices, access to health information, recognition of warning symptoms and early healthcare-seeking behavior. This finding supports the need for prevention strategies adapted to the population’s level of health literacy.

Marital status was also associated with case status. Participants who were married or living with a partner represented 80.0% of cases compared with 53.0% of controls, with a significant association in multivariate analysis (aOR = 3.72; 95% CI: 1.19–11.66; p = 0.024). However, this association should remain secondary in interpretation, as marital status is not an established causal factor for colorectal cancer. It may reflect differences in age, family structure, dietary habits or healthcare-seeking behavior.

The anthropometric findings were mixed. Overweight or obesity according to BMI was less frequent among cases than controls, 34.0% versus 63.0%, respectively. In bivariate analysis, this category was inversely associated with case status (crude OR = 0.30; 95% CI: 0.14–0.63; p = 0.009). Conversely, abdominal obesity was more frequent among cases than controls, 44.0% and 27.0%, respectively, and was significantly associated with case status in bivariate analysis (crude OR = 2.12; 95% CI: 1.04–4.33; p = 0.036). After adjustment, this association did not remain significant in the multivariate model (aOR = 1.77; 95% CI: 0.50–6.31; p = 0.377).

These findings partially differ from regional studies. In Uganda, Wismayer et al. reported in 2024, in a case–control study including 129 colorectal cancer cases and 258 controls, that obesity was more frequent among cases than controls, with 14.7% of cases having a BMI ≥ 30 kg/m² compared with 8.5% of controls. Obesity was associated with colorectal cancer in crude analysis (OR = 2.37; 95% CI: 1.21–4.64), but this association did not persist after adjustment (aOR = 1.23; 95% CI: 0.44–3.41) [5]. In Algeria, Negrichi and Taleb observed, among 64 cases and 64 controls, a higher frequency of obesity among cases than controls, 37.5% versus 20.3%, with a significant association (p = 0.048) [6]. In Morocco, the systematic review by Amsdar et al. also reported that some Moroccan studies found a higher risk of colorectal cancer among overweight or obese individuals, although findings varied across studies [9].

At the international level, the association between adiposity and colorectal cancer is well documented. The WCRF/AICR considers excess body fatness an established risk factor for colorectal cancer [10]. Similarly, Lauby-Secretan et al., in the 2016 IARC Working Group report, concluded that excess body fatness is associated with several cancers, including colorectal cancer [3]. Meta-analyses by Moghaddam et al. and Ma et al. also confirmed a positive association between general obesity and colorectal cancer risk [11,12].

The inverse association observed between high BMI and case status in the present study is likely related to reverse causality. Patients with colorectal cancer may lose weight before or after diagnosis, making BMI measured at the time of the survey a poor indicator of pre-disease body size. Nevertheless, the bivariate association with abdominal obesity remains biologically plausible, as central adiposity more accurately reflects metabolic and inflammatory disturbances related to colorectal cancer risk. In Nouakchott, future studies should document usual pre-disease weight, recent weight change, waist circumference and waist-to-hip ratio.

No behavioral factor was significantly associated with colorectal cancer. Smoking was reported by 10.0% of cases and 11.0% of controls, with no significant association (OR = 0.90; 95% CI: 0.29–2.80; p = 0.86). This contrasts with Yang et al., who reported in China in 2021 an association between smoking and left-sided colorectal cancer (aOR = 1.25; 95% CI: 1.16–1.34) [13], and with GBD 2019 data highlighting the global burden of smoking-related diseases [14]. In the present study, this null finding may reflect the low reported prevalence of smoking, possible underreporting, or limited statistical power.

Physical activity was also not associated with colorectal cancer. Lack of physical activity was reported by 64.0% of cases and 57.0% of controls (OR = 1.34; 95% CI: 0.67–2.68; p = 0.39). This finding differs from Moore et al., who showed in 2016, in a study including 1.44 million adults, that high leisure-time physical activity was associated with a lower risk of colon cancer (HR = 0.84; 95% CI: 0.77–0.91) [15]. In Arab countries, Sharara et al. also emphasized in 2018 the importance of physical inactivity as a public health issue [16]. The absence of association in the present study may be due to the simplified measurement of physical activity, which did not account for duration, intensity or type of activity.

Eating outside the home was not associated with colorectal cancer (60.0% among cases versus 61.0% among controls; OR = 0.96; 95% CI: 0.48–1.91; p = 0.91). This differs from Bouzini et al. in Morocco, who reported that fast-food consumption more than three times per week was strongly associated with early-onset colorectal cancer (aOR = 9.17; 95% CI: 2.93–28.57) [17]. This discrepancy is likely explained by the fact that the variable “eating outside the home” did not distinguish fast food from traditional or family meals.

Occupational exposure and sun exposure were not significantly associated with colorectal cancer. Occupational exposure was reported by 10.0% of cases and 9.0% of controls, while sun exposure at least once per week was reported by 30.0% of cases and 35.0% of controls. Regarding sun exposure, Hernández-Alonso et al. reported that higher circulating vitamin D levels were associated with reduced colorectal cancer risk (OR = 0.61; 95% CI: 0.52–0.71) [18], but the present study did not include biological measurement of vitamin D.

The absence of reported alcohol consumption in the present study population is consistent with the religious and sociocultural context of Mauritania, a predominantly Muslim country where alcohol consumption is generally prohibited. This finding, while limiting the statistical analysis of alcohol as a risk factor, reflects a genuine population characteristic and may partly explain a distinct colorectal cancer risk profile compared to Western populations.

In the present study, the main dietary findings concerned salty, canned, processed and sugary foods. High consumption of salty foods was more frequent among cases than controls (80.0% vs. 55.0%) and remained associated with case status after adjustment. However, the very wide confidence interval indicates substantial statistical imprecision; this result should therefore be interpreted as a signal of an unfavorable salty and processed dietary pattern rather than as evidence of an isolated causal effect of salt. High consumption of canned foods was also more frequent among cases (60.0% vs. 35.0%; OR = 2.79; 95% CI: 1.37–5.66; p = 0.004), but did not persist in the multivariate model. These findings suggest a broader processed and salty dietary pattern rather than an isolated effect of salt. They are consistent with data from Wismayer et al. in Uganda, 2024, and Amsdar et al. in Morocco, 2025, where modern processed meats were associated with colorectal cancer (aOR = 3.19; 95% CI: 2.06–4.92) [5,19]. They also align with the Moroccan review by Amsdar et al., 2024, and the Gulf countries review by Adam et al., 2024, which reported increased risk with diets rich in processed foods, red meat and refined carbohydrates [9,20]. Internationally, the WCRF/AICR considers processed meat a convincing cause of colorectal cancer, with an estimated 18% increase in risk per 50 g/day [10]. High red meat consumption was common in both groups and was not significantly associated with colorectal cancer (76.0% among cases vs. 71.0% among controls; OR = 1.29; p = 0.53). Processed meat showed a clearer trend toward association (20.0% vs. 9.0%; OR = 2.53; p = 0.06). This trend is consistent with Azzeh et al. in Saudi Arabia, 2017, and Hur et al., 2019, who reported an association between red meat, processed meat and colorectal cancer risk [7,21]. The lack of statistical significance for red meat in the present study may be explained by high consumption in both groups, the absence of portion quantification and the limited sample size. Foods usually considered protective showed expected but non-significant trends. Low fruit intake was reported by 70.0% of cases compared with 55.0% of controls (OR = 1.91; p = 0.07), low dairy intake by 44.0% versus 30.0% (OR = 1.83; p = 0.09), and low vegetable intake by 40.0% versus 35.0% (OR = 1.24; p = 0.54). These trends are consistent with Amsdar et al. in Morocco, 2025, who reported a protective effect of vegetables (aOR = 0.32; 95% CI: 0.20–0.51) and dairy products (aOR = 0.62; 95% CI: 0.39–0.97) [19]. They also align with Aune et al., 2011, on the beneficial effect of dietary fiber and whole grains, and with WCRF/AICR recommendations promoting fruits, vegetables, legumes and whole grains [10,22,23].

High consumption of refined sugars was significantly more frequent among cases than controls (90.0% vs. 63.0%; OR = 5.29; 95% CI: 1.93–14.49; p = 0.001). This finding is consistent with Kanehara et al., 2024, and Makarem et al., 2018, who reported associations between high sugar intake, high glycemic load and colorectal cancer risk [24,25]. Conversely, high consumption of sugar-sweetened beverages was less frequent among cases than controls (4.0% vs. 32.0%) and was inversely associated with case status. This finding should be interpreted cautiously, as international evidence remains heterogeneous: Hauan et al. found no substantial association in the Norwegian NOWAC cohort, whereas Feng et al. and Jatho et al. suggested a possible association between sugar-sweetened beverages and colorectal or gastrointestinal cancer risk [26–28]. In the present study, this inverse association may reflect reverse causality, dietary changes after diagnosis or underreporting.

Regarding tea, no significant association was observed with colorectal cancer, whereas coffee consumption was markedly less frequent among cases than controls (4.0% vs. 45.0%) and remained inversely associated after adjustment. International evidence remains mixed: Um et al., 2020, found no significant association, whereas Poole et al., 2017, reported a moderate inverse association [29,30]. In the present study, this finding should remain exploratory, as only two cases reported coffee consumption. Overall, these findings support nutrition-related prevention interventions in Nouakchott focused on reducing salty, processed and sugary foods, while promoting fiber-rich foods, fruits, vegetables, legumes and minimally processed foods. This study provides original data on factors associated with colorectal cancer in Nouakchott, in a context where local evidence remains limited. Its case–control design, with two controls per case, the use of a standardized questionnaire, and the combined assessment of sociodemographic, dietary, behavioral and anthropometric factors represent its main strengths. However, some limitations should be acknowledged. The sample size was limited, which reduced statistical power and resulted in wide confidence intervals for some estimates. The case–control design may have introduced recall bias and reverse causality, particularly for dietary habits that may have changed after diagnosis. Furthermore, the logistic regression model included eight variables for 50 cases, which does not satisfy the recommended rule of 10 events per variable (EPV ≥10). This may have led to model instability and overfitting, particularly for the estimate related to salty food consumption. The odds ratio for this variable (aOR = 47.45) should therefore be interpreted as exploratory. Future studies with larger sample sizes are needed to obtain more stable and precise estimates.

Therefore, the observed associations should be interpreted as epidemiological signals requiring confirmation through larger studies, ideally multicenter or prospective.

## 5 Conclusion

This case–control study conducted in Nouakchott highlights the potential role of social determinants and certain dietary habits in the profile of colorectal cancer cases. After adjustment, low educational level and high consumption of salty foods were the most defensible associations. Being married or living with a partner was also associated with case status, although this finding should be interpreted with caution.

The findings suggest a dietary profile characterized by high consumption of salty, canned, processed and refined sugar-rich foods. The inverse associations observed for coffee and sugar-sweetened beverages should be considered exploratory, given the potential for reverse causality, reporting bias and the small number of exposed cases.

These results support the strengthening of nutrition-related prevention and health education interventions in Nouakchott, particularly among populations with low educational attainment. Larger studies with more precise assessment of dietary and behavioral exposures are needed to confirm these associations and guide colorectal cancer prevention strategies in Mauritania.

These findings may apply mainly to adults in urban settings undergoing nutritional transition and may not be directly generalizable to rural Mauritanian populations.

## Data Availability

The anonymized dataset supporting the findings of this study is publicly available on Zenodo at: https://doi.org/10.5281/zenodo.20310085

## 6 Declarations

### Ethics approval and consent to participate

This study was conducted in accordance with the ethical principles governing research involving human participants and in line with the Declaration of Helsinki. The study protocol received ethical approval from the Health Research Ethics Committee of the Ministry of Health of Mauritania, under reference number 047-2026/MS/CNERS, dated January 22, 2026. Administrative authorization was also obtained from the relevant institutions, particularly the National Oncology Center. Oral informed consent was obtained from all participants before inclusion.

### Consent for publication

Not applicable. This manuscript does not contain any individual person’s data, images or clinical details.

## Competing interests

The authors declare that they have no competing interests.

## Funding

This research received no specific grant from any funding agency in the public, commercial or not-for-profit sectors.

## Authors’ contributions

NT conceived the study, developed the study protocol, coordinated data collection, performed data management and statistical analysis, interpreted the results, and drafted the manuscript. All co-authors contributed to methodological and scientific supervision, interpretation of findings, and critical revision of the manuscript. All authors read and approved the final manuscript.

## Use of artificial intelligence tools

Artificial intelligence-assisted tools were used for language editing and academic style improvement. All AI-assisted outputs were reviewed and verified by the authors, who take full responsibility for the content, accuracy, interpretation and integrity of the manuscript. No AI tool was used for data collection, statistical analysis or generation of original research data.

## Acknowledgements

The authors would like to thank the Health Research Ethics Committee of the Ministry of Health of Mauritania, the National Oncology Center, the data collection team, and all participants who agreed to take part in this study.

## Notes

### Competing Interest Statement

The authors have declared no competing interest.

### Funding Statement

The author(s) received no specific funding for this work.

### Author Declarations

Health Research Ethics Committee of the Ministry of Health of Mauritania. Approval reference: 047-2026/MS/CNERS, dated January 22, 2026.

